# Social contact and inequalities in depression and loneliness among older adults: A mediation analysis of the English Longitudinal Study of Ageing

**DOI:** 10.1101/2020.07.01.20143990

**Authors:** Michael J Green, Elise Whitley, Claire L Niedzwiedz, Richard J Shaw, S Vittal Katikireddi

## Abstract

**Background:** Social contact, including remote contact (by telephone, email, letter or text), could help reduce social inequalities in depression and loneliness among older adults.

**Methods:** Data were from the 8^th^ wave of the English Longitudinal Study of Aging (2016/17), stratified by age (n=1,635 aged <65; n=4,123 aged 65+). Inverse probability weighting was used to estimate average effects of weekly in-person and remote social contact on depression (score of 3+ on 8-item CES-D scale) and two measures of loneliness (sometimes/often feels lonely vs hardly ever/never; and top quintile of UCLA loneliness scale vs all others). We also estimated controlled direct effects of education, partner status, and wealth on loneliness and depression under two scenarios: 1) universal infrequent (<weekly) in-person social contact; and 2) universal weekly remote social contact.

**Results:** Weekly in-person social contact was associated with reduced odds of depression and loneliness, but associations with remote social contact were weak. Lower education raised odds of depression and loneliness, but differences were attenuated with infrequent in-person contact. Respondents living alone experienced more depression and loneliness than those living with a partner, and less wealth was associated with more depression. With universal infrequent in-person contact, these differences narrowed among those aged under 65 but widened among those aged 65+. Universal weekly remote contact had little impact on inequalities.

**Conclusions:** Reduced in-person social contact may increase depression and loneliness among older adults, especially for those aged 65+ who live alone. Reliance on remote social contact seems unlikely to compensate for social inequalities.

## 1. Introduction

### 1.1 Background

Many older adults experience depression and loneliness (Age UK, 2011; Green & Benzeval, 2011; Hansen & Slagsvold, 2016; Meeks et al., 2011; Niedzwiedz et al., 2016), which are associated with lower quality of life, health-risk behaviours and poor physical health (Beutel et al., 2017; Courtin & Knapp, 2017; Eaton et al., 2008; Meeks et al., 2011; Rodda et al., 2011; Shankar et al., 2011; Valtorta et al., 2016), and are socially patterned, being more common among older adults from poorer socioeconomic backgrounds and who live alone (Age UK, 2011; Beutel et al., 2017; Chang-Quan et al., 2010; Green & Benzeval, 2011; Hansen & Slagsvold, 2016; Kamiya et al., 2013; Meeks et al., 2011; Niedzwiedz et al., 2016). Social mitigation responses to the worldwide Covid-19 pandemic, emphasising physical distance from others, may exacerbate these issues (Armitage & Nellums, 2020; Douglas et al., 2020; Holmes et al., 2020).

Social contact with friends and family may contribute to inequalities in loneliness and depression. Social contact may be in-person or remote (i.e. via phone/internet etc.), but is distinct from loneliness, which is a perceived feeling of social isolation (Age UK, 2011; Courtin & Knapp, 2017; Hughes et al., 2004; Niedzwiedz et al., 2016). One can feel lonely amid frequent social contacts, or not feel lonely with very few social contacts. Nevertheless, more frequent social contact is associated with less depression and loneliness (Age UK, 2011; Kearns et al., 2015; Niedzwiedz et al., 2016; Teo et al., 2015), and is more common among older people who are socioeconomically advantaged or who live with a partner (Ajrouch et al., 2005; Gray, 2009; Kearns et al., 2015).

The importance of digital communications has been increasing for some time, but remote contact has become especially salient during the covid-19 pandemic. Many countries have enacted ‘lockdown’ social mitigation measures, where reduced physical proximity to others is intended to slow the infection transmission rate, but such measures are likely to have psychological impacts (Armitage & Nellums, 2020; Brooks et al., 2020; Douglas et al., 2020; Sood, 2020). Identified as high risk for covid-19, older adults have been advised to follow stringent social distancing measures to avoid infection (Armitage & Nellums, 2020; Douglas et al., 2020). Maintaining or increasing remote social contact has been promoted to mitigate impacts of social distancing (Armitage & Nellums, 2020; Brooks et al., 2020; Chatterjee & Yatnatti, 2020; Sood, 2020), but remote contact has been less strongly associated with depression and loneliness than in-person contact (Teo et al., 2015), so may not compensate adequately. Reducing in-person social contact or increasing remote social contact could both potentially narrow inequalities (by reducing social patterning of salutary factors) or widen them (if some benefit more from contact than others) (Niedzwiedz et al., 2016).

Understanding the different contributions that in-person and remote social contacts make to inequalities in loneliness and depression is important in informing policy responses to improve health, especially with regards to mitigation of social distancing.

### 1.2 Research Questions

We use a mediation analysis of the most recent wave of data from the English Longitudinal Study of Aging, to estimate answers to the following questions:

1. What are the effects of in-person and remote social contact on loneliness and depression?
2. To what extent are inequalities in loneliness and depression affected by making in-person social contact infrequent for everyone?
3. To what extent are inequalities in loneliness and depression affected by making remote social contact frequent for everyone?

## 2. Methods

### 2.1 Sample

The English Longitudinal Study of Ageing (ELSA) is a large-scale, representative, longitudinal panel study of people aged 50 and over living in private households in England (Marmot et al., 2018). The core sample has been drawn from respondents to the Health Survey for England (HSE) since 1998. We focus on 7,223 core sample members who were interviewed at the most recent survey wave (wave 8: 2016/2017; 82.4% of those who were eligible for inclusion because they were still alive and living in the UK). We excluded 930 respondents who did not return a self-completion questionnaire, and a further 535 respondents with missing data on relevant variables (see Figure 1). This resulted in a final sample of 5,758 respondents (79.7% of the core sample members interviewed).

**Figure 1:**
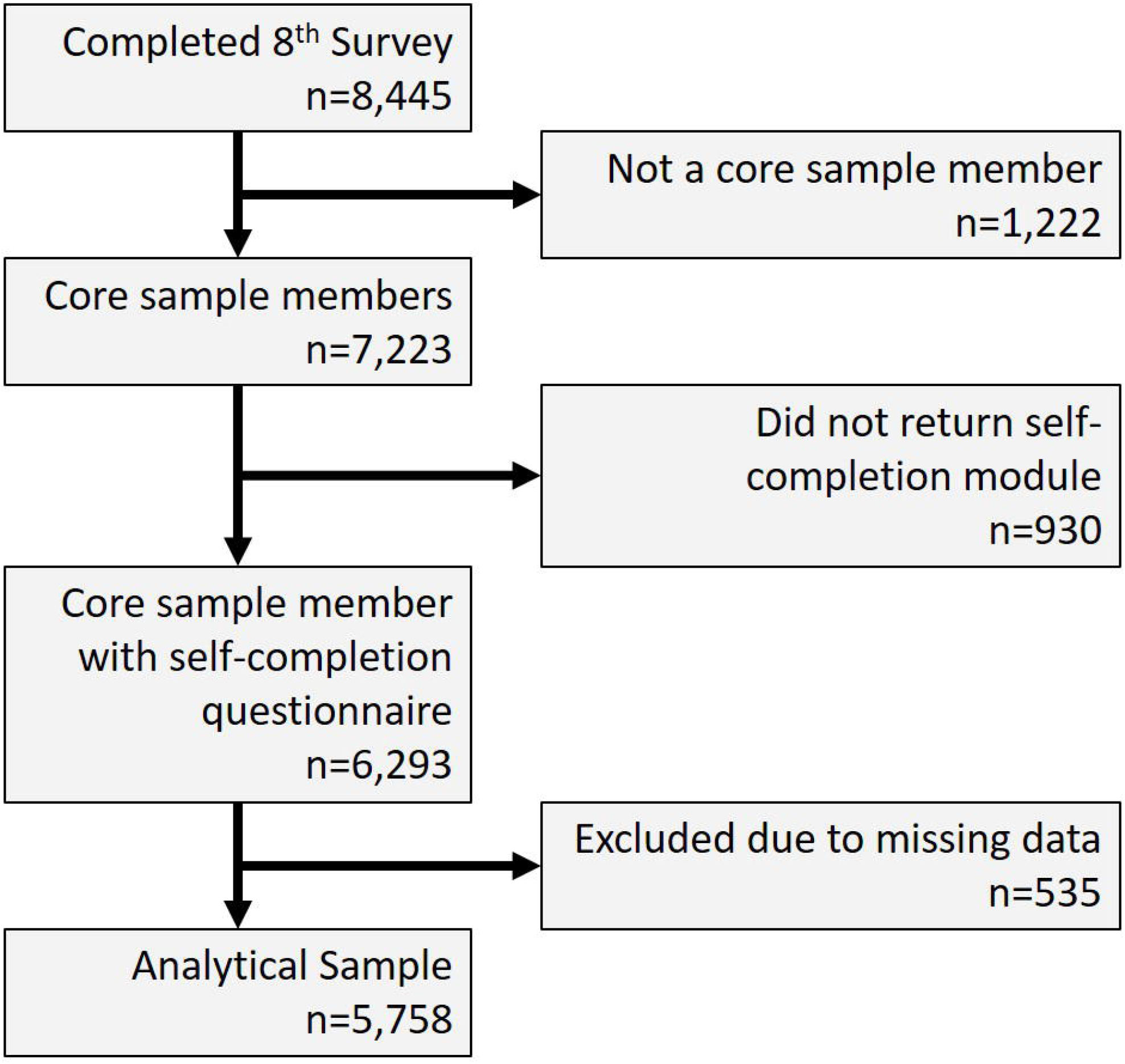
Derivation of analytical sample.

### 2.2 Measures

#### 2.2.1 Loneliness and depression

Depression was based on an 8-item version of the Centre for Epidemiologic Studies Depression Scale (CES-D; Radloff, 1977), with scores ranging from 0 (least depressed) to 8 (most depressed). Scores of 3+ were coded as probable psychiatric cases, a threshold previously validated against standardised psychiatric interviews in older populations (Turvey et al., 1999). Loneliness at wave 8 was assessed in two ways. Firstly, we considered responses to the self-complete questionnaire question “How often do you feel lonely?” (some of the time/often versus hardly ever/never). Secondly we used the 3-item UCLA loneliness scale (Hughes et al., 2004), which incorporates responses to three questions: “How often do you feel that you lack companionship?”; “How often do you feel left out?”; and “How often do you feel isolated from others?”. Scores ranged between 3 and 9 and we compared the loneliest quintile (scores 6-9) versus the other four (scores 3-5).

#### 2.2.3 Primary exposures

We considered three potential sources of inequality in loneliness and depression: 1) education, compared respondents with A level equivalent or higher qualifications (i.e. beyond completion of ordinary schooling) versus those with lesser or no qualifications; 2) partner status, comparing those living with a partner (regardless of marital status) versus those not living with a partner; and 3) wealth, measured using quintiles of net total non-pension household wealth (a summary of the total value of the financial, physical and housing wealth owned by the household; derivation of this variable has been described in full elsewhere) (Demakakos et al., 2016). For wealth, we focus on results comparing the least to the wealthiest quintile (with results for other quintiles included as supplementary information).

#### 2.2.2 Social contact

Respondents were asked how often, on average, they met up with their children, other family members or friends (separate questions for each), and we compared those meeting up with at least one of their children, other family members or friends at least weekly versus those meeting up less frequently or reporting no children, family or friends. Weekly remote contact was coded similarly using questions on frequency of contact by telephone, letter, email or text with children, other family or friends (again separate questions were asked for each).

#### 2.2.3 Confounding variables

Confounding variables included: sex, five-year age group, ethnicity (white versus non-white), government office region, any children in household (yes/no), any children outside household (yes/no), number of people in household other than respondent and partner, whether respondent felt close to their children, other family or friends, housing tenure (owner-occupant/rented), economic activity (in paid employment/not), social class (I/II, III non-manual, III manual, IV/V) of last known occupation coded according to the British Registrar General’s scheme which indicates occupational differences in status and economic resources (Galobardes et al., 2006; Office of Population Censuses and Surveys, 1980), number of problems with instrumental activities of daily living (Lawton & Brody, 1969), and whether the respondent was a member of any clubs or organisations.

#### 2.2.4 Statistical Analyses

All models used weights to adjust for sampling and drop-out (Marmot et al., 2018). Preliminary analyses indicated interactions with age, so we stratified our analyses between those aged under 65 (the UK retirement age) and those aged 65 or more.

For research question 1 we separately estimated the effects of in-person and remote social contact on loneliness and depression, using inverse probability weighting (IPW) (Austin, 2011). This is described more fully in the Appendix (section 5.1) but adjusts for measured potential confounders (including education, partner status and wealth). Estimates represent average effects of in-person and remote social contact on depression and loneliness within the sample (assuming no residual confounding or reverse causation).

For research questions 2 and 3, we assume social contact may mediate effects of either education, partner status or wealth on loneliness and depression outcomes, as shown in Figure 2. We distinguish pre- and post-exposure confounders. Pre-exposure confounders (X) are potential common causes of the exposure, the mediator and the outcome. In contrast, post-exposure confounders (C) are potential common causes of the mediator and the outcome but may (or may not) be caused by the exposure. This distinction is important for estimating the effect of the exposure after intervention on the mediator (VanderWeele, 2009). Table 1 shows which variables were considered as pre/post confounders depending on the exposure in question. Given ambiguity regarding causal direction between partner status and socioeconomic variables (housing tenure, social class, economic activity and wealth) we performed sensitivity analyses with these variables re-positioned as pre-exposure confounders.

**Table 1:** Analysis Variables.

| **Exposure (E)** | **Pre-Exposure Confounders (X)** | **Post-Exposure Confounders (C)** |
| --- | --- | --- |
| Education | Gender<br>Age<br>Ethnicity<br>Region | Remote/In-person social contact<sup>a</sup><br>Partner status<br>Wealth<br>Child in Household<br>Child out of Household<br>Other Adults in Household<br>Close to Children<br>Close to Family<br>Close to Friends<br>Housing Tenure<br>Economic Activity<br>Social Class<br>IADL<br>Member of social organisation |
| Partner status | Gender<br>Age<br>Ethnicity<br>Region<br>Education | Remote/In-person social contact<sup>a</sup><br>Wealth<sup>b</sup><br>Child in Household<br>Child out of Household<br>Other Adults in Household<br>Close to Children<br>Close to Family<br>Close to Friends<br>Housing Tenure<sup>b</sup><br>Economic Activity<sup>b</sup><br>Social Class<sup>b</sup><br>IADL<br>Member of social organisation |
| Wealth | Gender<br>Age<br>Ethnicity<br>Education<br>Partner status<br>Housing Tenure<br>Economic Activity<br>Social Class | Remote/In-person social contact<sup>a</sup><br>Child in Household<br>Child out of Household<br>Other Adults in Household<br>Close to Children<br>Close to Family<br>Close to Friends<br>IADL<br>Member of a social organisation |

<sup>a</sup>Depending on whether in-person or remote social contact was the mediator of interest, the other was included as a post-exposure confounder.
<sup>b</sup>Since the causal direction between partner status and these socioeconomic variables was particularly ambiguous we performed sensitivity analyses where these variables were re-positioned as pre-exposure confounders.
IADL=Instrumental Activities of Daily Living.

**Figure 2:**
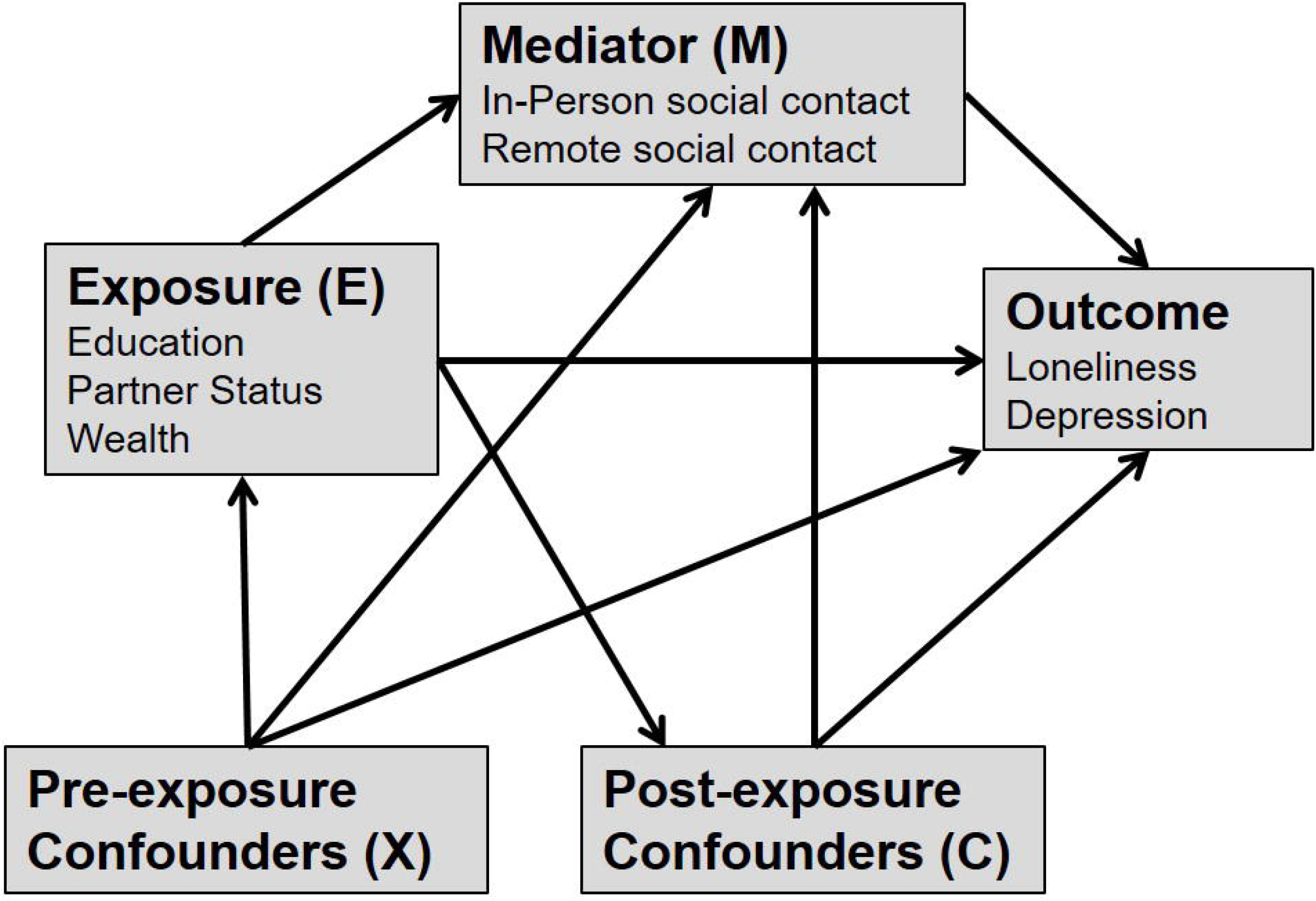
Assumptions about causal direction in our analyses.

Total exposure effects were estimated using IPW (see Appendix 5.2) to estimate the average effects of education, partner status or wealth on depression and loneliness, adjusting for pre-exposure confounders. These were compared to estimates of controlled direct effects (CDEs) for each exposure on loneliness and depression, which were estimated using inverse-probability weighted marginal structural models to adjust appropriately for pre- and post-exposure confounders (see Appendix 5.3; VanderWeele, 2009). The CDE represents the estimated effect of the exposure under a hypothetical intervention to set the mediator (social contact) to the same value for all respondents. Estimation allows for an interaction between the exposure and social contact (e.g. if social contact is more beneficial for some groups than others), such that CDEs can differ depending on the level that social contact is set to. We estimated CDEs for two scenarios: 1) universal infrequent (<weekly) in-person social contact; and 2) universal weekly remote social contact.

## 3. Results

### 3.1 Sample characteristics

Supplementary Table 1 presents characteristics of the analytical sample by age. Respondents aged 65 or more (n=4,123) were more likely to be white, living alone, less educated, and have frequent in-person social contact than those aged under 65 (n=1,635). Supplementary Table 1 also summarises characteristics of those who were excluded due to missing data, who were more likely to be at the extremes of the age range, from ethnic minority groups, living with a partner, less educated, less wealthy, have less frequent in-person and remote social contact, and were more likely to feel lonely or depressed.

### 3.2 Frequency of Social Contact

Table 2 shows proportions within each age group experiencing less than weekly in-person and remote social contact. Infrequent in-person social contact was more common in both age groups than infrequent remote social contact. Infrequent in-person social contact was most common among those with more education, who lived with a partner, or were wealthier. These differences were more pronounced in the 65+ age group, and the wealth difference was in the opposite direction for the under 65 age group. There was little social patterning of remote social contact by education, and the direction of patterning by partner status differed by age. Infrequent remote contact was more common in the lowest than the highest wealth quintile in the under 65 age group.

**Table 2:** Proportions experiencing infrequent in-person and remote social contact by education, partner status and wealth.

|  |  | <Weekly in-person social contact (%) |  | <Weekly remote social contact (%) |  |
| --- | --- | --- | --- | --- | --- |
|  |  | *Age <65* | *Age 65+* | *Age <65* | *Age 65+* |
| **Education** |  |  |  |  |  |
|  | Low | 26.0 | 22.1 | 10.9 | 10.4 |
|  | High | 29.3 | 27.7 | 11.9 | 11.8 |
| **Partner status** |  |  |  |  |  |
|  | Lives alone | 26.9 | 17.4 | 15.4 | 8.7 |
|  | With partner | 27.8 | 27.8 | 9.9 | 12.2 |
| **Wealth** |  |  |  |  |  |
|  | Lowest Quintile | 28.8 | 20.3 | 16.8 | 11.1 |
|  | Highest Quintile | 26.8 | 29.5 | 7.4 | 10.0 |
| **Total** |  | 27.5 | 24.3 | 11.4 | 11.0 |

### 3.3 Estimated Effects of Social Contact

Table 3 shows estimates of the average effects of in-person and remote social contact on loneliness and depression. Weekly in-person social contact was associated with reduced odds of loneliness in both age groups, even after adjustment for confounders. Adjustment also revealed a borderline association between more frequent in-person social contact and reduced odds of depression in the 65+ age group. There were no clear associations with remote social contact after adjustment for confounders. Findings using continuous scores for depression and loneliness were similar (Supplementary Table 2).

**Table 3:** Estimates of effects of in-person and remote social contact on depression and loneliness.

|  | Weekly in-person social contact<br>(vs. less than weekly) |  |  |  | Weekly remote social contact<br>(vs. less than weekly) |  |  |  |
| --- | --- | --- | --- | --- | --- | --- | --- | --- |
|  | Age <65<br>N=1,635 |  | Age 65+<br>N=4,123 |  | Age <65<br>N=1,635 |  | Age 65+<br>N=4,123 |  |
|  | OR | 95% CI | OR | 95% CI | OR | 95% CI | OR | 95% CI |
| *CES-D Depression* |  |  |  |  |  |  |  |  |
| Sample weighted association | 0.78 | 0.54-1.13 | 0.99 | 0.81-1.21 | 0.57 | 0.36-0.91 | 1.16 | 0.86-1.57 |
| ATE estimate<sup>a</sup> | 0.96 | 0.63-1.47 | 0.80 | 0.63-1.01 | 0.83 | 0.39-1.79 | 1.29 | 0.81-2.04 |
| *Sometimes/Often Feels Lonely* |  |  |  |  |  |  |  |  |
| Sample weighted association | 0.73 | 0.54-1.00 | 0.88 | 0.74-1.05 | 0.87 | 0.56-1.34 | 0.86 | 0.68-1.09 |
| ATE estimate<sup>a</sup> | 0.73 | 0.52-1.01 | 0.75 | 0.62-0.91 | 1.36 | 0.69-2.67 | 0.94 | 0.66-1.33 |
| *UCLA Loneliness* |  |  |  |  |  |  |  |  |
| Sample weighted association | 0.59 | 0.42-0.85 | 0.68 | 0.57-0.83 | 0.57 | 0.36-0.90 | 0.67 | 0.51-0.87 |
| ATE estimate<sup>a</sup> | 0.67 | 0.46-0.99 | 0.61 | 0.49-0.76 | 1.07 | 0.50-2.29 | 0.78 | 0.53-1.15 |

<sup>a</sup>ATE: Average treatment effect, i.e. the estimated average effect of weekly social contact within the sample after adjusting for education, partner status, wealth and all other confounders listed in Table 1. These estimates assume no residual confounding or reverse causation.

### 3.4 Estimated Effects of Education, Partner Status and Wealth

Table 4 shows ATE and CDE estimates of education, partner status and wealth on depression and loneliness for those both below and over the age of 65. Findings were similar with continuous measures of depression and loneliness (Supplementary Table 3). Lower education was associated with more loneliness and depression in both age groups, and the CDE estimates suggested that universally infrequent in-person social contact would attenuate this inequality. CDE estimates of ensuring weekly remote contact for everyone indicated relatively little impact on inequalities in loneliness, but some minor attenuation of inequalities in depression.

**Table 4:**
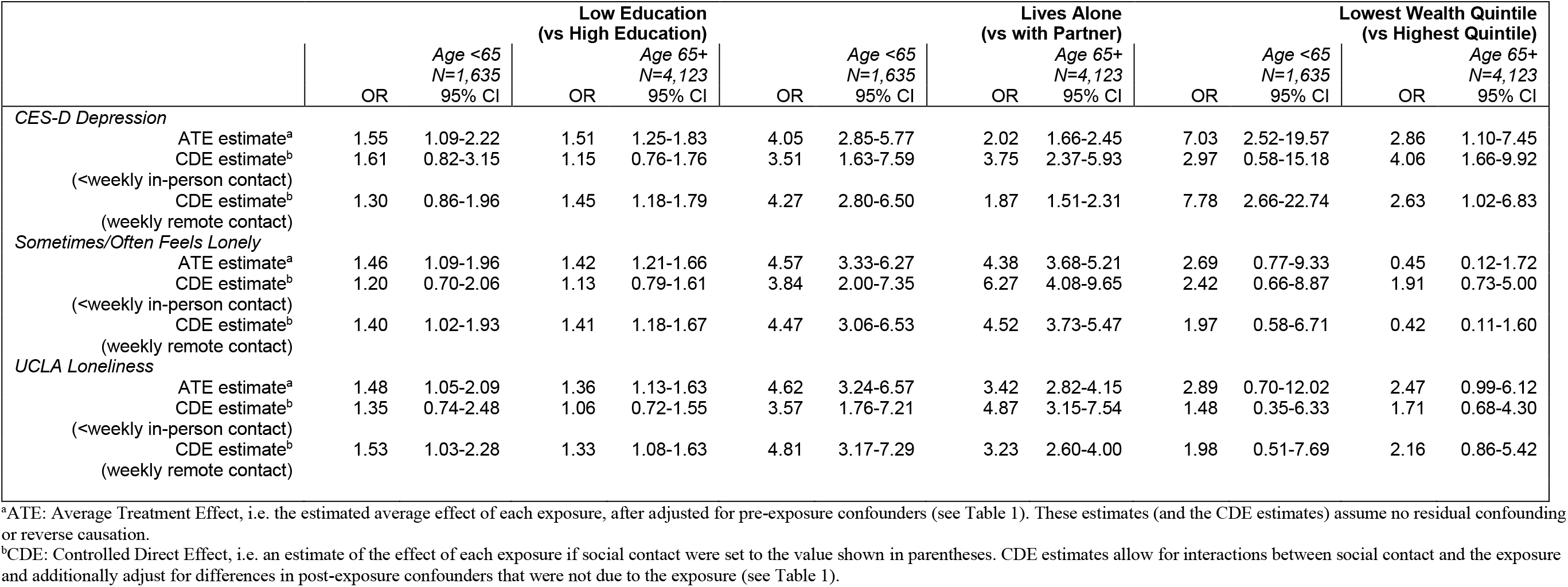
Estimates for effects of education, partner status and wealth on depression and loneliness.

Living alone was associated with more depression and loneliness than living with a partner. CDE estimates for universal infrequent in-person social contact indicated that these differences would widen among those aged over 65 but narrow among those aged under 65. CDE estimates for ensuring weekly remote contact for everyone indicated relatively little impact on these inequalities. Findings for partner status were similar with socioeconomic confounders re-positioned as pre-exposure confounders (Supplementary Table 4).

Being in the least wealthy compared to the wealthiest quintile was not clearly associated with loneliness except for the UCLA loneliness measure among the 65+ age group but was associated with greater odds of depression in both age groups. CDE estimates for universal infrequent in-person social contact indicated attenuation of this inequality in depression among those aged under 65 and the inequality in loneliness among those aged 65+, but increased inequalities in depression among those aged 65+. CDE estimates of ensuring weekly remote contact for everyone indicated relatively little impact on these inequalities, though there was some attenuation among the 65+ age group. Results for other wealth quintiles (Supplementary Table 5) were similar, though there was not a clear linear wealth gradient.

## 4. Discussion

### 4.1 Summary of Findings

In-person social contact was associated with reduced risk of depression and loneliness in older adults in England, while associations with remote social contact were relatively weak. We estimated effects of restricting everyone to infrequent (<weekly) in-person social contact and findings suggested narrower inequalities in depression and loneliness by education. Inequalities in depression and loneliness by partner status and in depression by wealth were impacted differently depending on age, with restricted in-person contact narrowing these inequalities in those aged <65 but widening them among adults aged 65+. Estimates for ensuring everyone received weekly remote contact indicated relatively little impact on inequalities.

### 4.2 Implications

Our findings are consistent with others showing associations between social contact and depression and loneliness (Age UK, 2011; Kearns et al., 2015; Niedzwiedz et al., 2016), with in-person contact more important than remote (Teo et al., 2015). While others found social contacts concentrated among the more advantaged (Ajrouch et al., 2005; Gray, 2009; Kearns et al., 2015), we found weekly in-person contact was less common among more advantaged adults, especially those aged 65+. Considering this and that in-person social contact was on average associated with less loneliness and depression, the narrowing of inequalities indicated by our CDE estimates (which allow for interaction between social contact and education) suggests heterogeneity in the impact of in-person social contact, with those of higher education deriving greater benefit. While we did adjust for close relationships with friends and other family members, this could be due to differences in the quality of contacts, or differences in frequency of contact beyond the weekly threshold used.

Our findings suggest that reductions in in-person social contact, e.g. under social mitigation measures related to covid-19, are likely to lead to increases in depression and loneliness in older adults. However, impacts may be experienced unequally and we estimated that those aged 65+ who live alone were particularly at risk for depression and loneliness under restrictions to in-person contact. This has been anticipated (Armitage & Nellums, 2020; Douglas et al., 2020), with remote social contact advised to mitigate these impacts (Brooks et al., 2020; Chatterjee & Yatnatti, 2020; Sood, 2020). We found that remote contact, at least as experienced pre-covid-19, seemed unlikely to compensate, with little effect on depression or loneliness, or on inequalities in depression and loneliness by education, partner status or wealth (even when assuming no disparities in establishing regular remote contact).

### 4.3 Limitations

Our estimates based on pre-covid-19 data may not necessarily generalise. For example, infrequent social contact under covid-19 mitigation measures, may have different effects to infrequent social contact experienced under other circumstances (Fancourt & Steptoe, 2020). Moreover, the covid-19 situation may have prompted qualitative improvements in remote contact that alter its effect. For example, our measure of remote contact did not specifically ask about contact via video-calling, which is likely to have become increasingly relevant during the pandemic (though respondents may have included this as telephone contact). A number of suggestions for helping older adults manage social isolation focus on enhancing benefits from remote contacts, such as cognitive-behavioral interventions and techniques (Van Orden et al., 2020), or interventions to set up remote ‘telehealth’ groups (Zubatsky et al., 2020). Our results, which estimate effects of remote contact as it was experienced pre-covid-19, should not be taken to mean that such efforts are likely to be ineffective, but highlight the importance of better understanding how benefit can be derived from remote contact, and how effective forms of remote contact might impact on inequalities (both issues likely to remain salient after the pandemic).

Additionally, we focus solely on mechanisms of social contact but there are several other mechanisms related to social mitigation of covid-19 that could impact inequalities in depression and loneliness among older adults, including bereavement, and anxieties or worries related to one’s health or economic situation or those of loved ones (Douglas et al., 2020). Furthermore, while data were cross-sectional, our analyses assumed causal direction from exposure to social contact to depression and loneliness. While we may be confident that education preceded other measures, our results could also be accounted for by social contacts affecting wealth or partner status, by depression or loneliness affecting social contact, or by residual confounding.

### 4.4 Conclusions

Reductions in in-person social contact could result in increased depression and loneliness among older adults. Adults aged 65+ who lived alone appeared especially vulnerable to reductions in social contact, with greater estimated increases in risk for depression and loneliness than similarly aged adults living with a partner. Inequalities in depression and loneliness by education could narrow as in-person contact is reduced, as those with more education seemed to derive more benefit from contact. Remote social contact seemed insufficient to mitigate adverse impacts or inequalities, and more attention is needed into how remote contact can become more efficacious, or to other strategies for addressing inequalities in depression and loneliness among older adults.

## Data Availability

Data were from the 8th wave of the English Longitudinal Study of Ageing and are available for use from the UK Data Service archive.

https://beta.ukdataservice.ac.uk/datacatalogue/studies/study?id=5050

## 5 Appendix

### 5.1 Estimating effects of social contact

Average Treatment Effects (ATEs) were estimated by regressing outcomes on each social contact variable, with each respondent assigned a weight equal to P(M)/P(M|E,X,C). M is their observed level of social contact. E represents the main exposure variables (education, partner status and wealth), which are confounders for the effect of social contact. X and C represent sets of pre and post-exposure confounders as listed in Table 1 (the pre/post distinction is not important here but is important for estimating impacts on inequalities below). The purpose of this weighting is to balance observed confounders (E, X and C) across levels of M. The probabilities required to calculate the weights were estimated via logistic regression models of M (with and without E, X and C).

### 5.2 Estimating total effects of exposure

Outcomes were regressed on each exposure (education, partner status, or wealth) with each respondent assigned a weight as P(E)/P(E|X), where E represents respondents’ observed values for the exposure in question, and X represents pre-exposure confounders as listed in Table 1. The purpose of this weighting is to balance the confounders (X) across levels of E. The probabilities required to calculate the weights were estimated via logistic regression models of E (with and without X).

### 5.3 Estimating controlled direct effects

Outcomes were regressed on the exposure (education, partner status or wealth), mediator (in-person or remote social contact), and an interaction term for the two factors. These marginal structural models were weighted using a combination of the weight for the total effect of the exposure (as described above; P(E)/P(E|X)) and a modified version of the social contact weight, this time calculated as P(M|E)/P(M|E,X,C). As the numerator of this weight is conditional on the exposure in question (E), it serves to balance intermediate confounders (C), but only within levels of the exposure, so differences in C that are due to the exposure are retained. The probabilities required to calculate these weights were estimated via logistic regression models of E and M (conditioned on E, X and/or C as required). This means estimates are adjusted for differences in pre-exposure confounders, and for differences in post-exposure confounders that are not due to the exposure. In contrast with traditional regression analyses, differences in post-exposure confounders that *are* due to the exposure are not adjusted out, and the indirect effect of the exposure on the outcome via C in Figure 2 is included.

## Acknowledgements

This work was supported by the UK Medical Research Council (grant numbers MC_UU_12017/13, MC_PC_17217, MR/R024774/1 to CN), the Scottish Government Chief Scientist Office (SPHSU13), and National Health Service Research Scotland (SCAF/15/02 to SVK). Funders had no role in conducting the study or preparing this article. The English Longitudinal Study of Ageing was developed by a team of researchers based at the University College London, NatCen Social Research, and the Institute for Fiscal Studies. The data were collected by NatCen Social Research. The funding is currently provided by the National Institute of Aging (R01AG017644), and a consortium of UK government departments coordinated by the National Institute for Health Research.

